# Localized high-risk prostate cancer harbors an androgen receptor low subpopulation susceptible to HER2 inhibition

**DOI:** 10.1101/2024.02.09.24302395

**Authors:** Scott Wilkinson, Anson T. Ku, Rosina T. Lis, Isaiah M. King, Daniel Low, Shana Y. Trostel, John R. Bright, Nicholas T. Terrigino, Anna Baj, John M. Fenimore, Chennan Li, BaoHan Vo, Caroline S. Jansen, Huihui Ye, Nichelle C. Whitlock, Stephanie A. Harmon, Nicole V. Carrabba, Rayann Atway, Ross Lake, Haydn T. Kissick, Peter A. Pinto, Peter L. Choyke, Baris Turkbey, William L. Dahut, Fatima Karzai, Adam G. Sowalsky

**Affiliations:** Genitourinary Malignancies Branch, National Cancer Institute, Bethesda, MD, USA; Department of Urology, Emory University School of Medicine, Atlanta, GA, USA; Department of Pathology and Department of Urology, University of California Los Angeles, Los Angeles, CA, USA; Molecular Imaging Branch, National Cancer Institute, Bethesda, MD, USA; Laboratory of Cancer Biology and Genetics, National Cancer Institute, Bethesda, MD, USA; Urologic Oncology Branch, National Cancer Institute, Bethesda, MD, USA

## Abstract

Patients diagnosed with localized high-risk prostate cancer have higher rates of recurrence, and the introduction of neoadjuvant intensive hormonal therapies seeks to treat occult micrometastatic disease by their addition to definitive treatment. Sufficient profiling of baseline disease has remained a challenge in enabling the in-depth assessment of phenotypes associated with exceptional vs. poor pathologic responses after treatment. In this study, we report comprehensive and integrative gene expression profiling of 37 locally advanced prostate tumors prior to six months of androgen deprivation therapy (ADT) plus the androgen receptor (AR) inhibitor enzalutamide prior to radical prostatectomy. A robust transcriptional program associated with HER2 activity was positively associated with poor outcome and opposed AR activity, even after adjusting for common genomic alterations in prostate cancer including *PTEN* loss and expression of the TMPRSS2:ERG fusion. Patients experiencing exceptional pathologic responses demonstrated lower levels of HER2 and phospho-HER2 by immunohistochemistry of biopsy tissues. The inverse correlation of AR and HER2 activity was found to be a universal feature of all aggressive prostate tumors, validated by transcriptional profiling an external cohort of 121 patients and immunostaining of tumors from 84 additional patients. Importantly, the AR activity-low, HER2 activity-high cells that resist ADT are a pre-existing subset of cells that can be targeted by HER2 inhibition alone or in combination with enzalutamide. In summary, we show that prostate tumors adopt an AR activity-low prior to antiandrogen exposure that can be exploited by treatment with HER2 inhibitors.

ClinicalTrials.gov registration: NCT02430480.

## INTRODUCTION

The molecular and clinical heterogeneity of localized high-risk (also known as locally advanced) prostate cancer (PCa) pose considerable obstacles for its diagnosis and treatment [1, 2]. Although the use of computed tomography (CT) and magnetic resonance imaging (MRI) has improved *in vivo* localization of extraprostatic disease that is associated with higher risk of prosate-specific antigen (PSA)/biochemical recurrence (BCR) or metastasis, significant weight is given to the volume and extent of dedifferentiation of PCa visualized on needle biopsies for assessing individualized risk [3]. Patients with locally advanced PCa receive multimodal therapies, which can include androgen deprivation therapy (ADT) with radiation therapy or adjuvant following surgery [3–6]. In patients treated by radical prostatectomy, adjuvant or neoadjuvant (before surgery) therapies function to reduce local tumor burden and target occult micrometastases that would eventually drive relapse [6, 7]. Phase 2 trials of different neoadjuvant therapies for localized high-risk PCa have tested multiple combinations of hormonal and chemohormonal therapies, including 2^nd^ and 3^rd^ generation androgen receptor (AR)-targeting agents [8–13], cytotoxic chemotherapy [14–16], and immunotherapy [17–19]. A persistent challenge common to all of these studies has been identifying robust baseline characteristics that would show long-term survival benefits based on prognostic features readily identified by short-term pathologic readouts.

In clinical trials of neoadjuvant intensive ADT, the biomarker showing the most robust performance towards predicting long-term outcome is residual cancer burden (RCB). Calculated based on the tumor cellularity of the final pathologic specimen, patients with lower RCB having complete pathologic responses (pCR) or minimal residual disease (MRD) demonstrated delayed BCR [20, 21], with three-year BCR-free rates of 95.2% (pCR/MRD) vs. 48.7% (greater than MRD). Using RCB or BCR as endpoints, molecular features detected in both posttreatment and baseline tissues have been reported with various prognostic potential, including the mutation status of *TP53* [22–25] and immunohistochemistry (IHC) against PTEN or ERG [9, 10, 22]. Across all reported studies, nearly every patient has shown at least some pathologic and/or imaging response to therapy. What remains unknown, however, is to what extent that clonal or subclonal intrinsic resistance to neoadjuvant intensive ADT, even in patients showing partial responses, can be targeted further.

Intrinsic resistance to targeted therapies involves cell-intrinsic mechanisms implicating the original target, which in the case of prostate cancer is the AR [26, 27]. In contrast to well-documented mechanisms of adaptive resistance involving AR point mutations or amplification after long-term exposure to ADT [27–29], a subset of aggressive prostate tumors display resistance to ADT as measured by transcriptional profiling of the tumor to have lower AR activity [24, 30, 31]. Due to the challenges of extensive profiling of biopsy tissues, genomic, histologic, and phenotypic properties of these low-AR tumors are not well characterized. Cellular mechanisms functioning to suppress AR activity, or which may otherwise be elevated in the absence of AR activity, have similarly remained elusive. Limited histologic and transcriptomic profiling of prostatectomy tissues after neoadjuvant ADT have identified potentially targetable “bypass” pathways, including AKT, HER2/HER3, CDK4/6, MET, BCL-2, and XBP1 [25, 32, 33]. Also proposed as an adaptive mechanism to prolonged ADT exposure, the detection of phosphorylated HER2 in metastatic PCa tumor biopsies supported previous clinical studies, which in this context showed little clinical benefit [32, 34–37]. Here, we present HER2 activity not as an adaptive bypass pathway but as an inherent and targetable mechanism of resistance in *de novo* locally advanced PCa with AR-low characteristics.

## RESULTS

### Prostate tumors exhibiting poor pathologic response to neoadjuvant ADT plus enzalutamide harbor a transcriptional signature of elevated HER2 activity

Using whole-transcriptome sequencing, we assessed patterns of gene expression and pathway activation in 147 tumor foci isolated from prostate biopsies prior to undergoing six months of intense neoadjuvant ADT (**Fig. 1A**). These foci were sampled using laser capture microdissection from 48 distinct MRI-visible lesions in 37 patients who participated in our clinical trial, with a median of two biopsy blocks per patient and a median of two foci per block sequenced. Most foci were isolated separately on the basis of differing histologic features, such as Gleason pattern or variability in key PCa markers such as PTEN or ERG staining.

**Figure 1.**
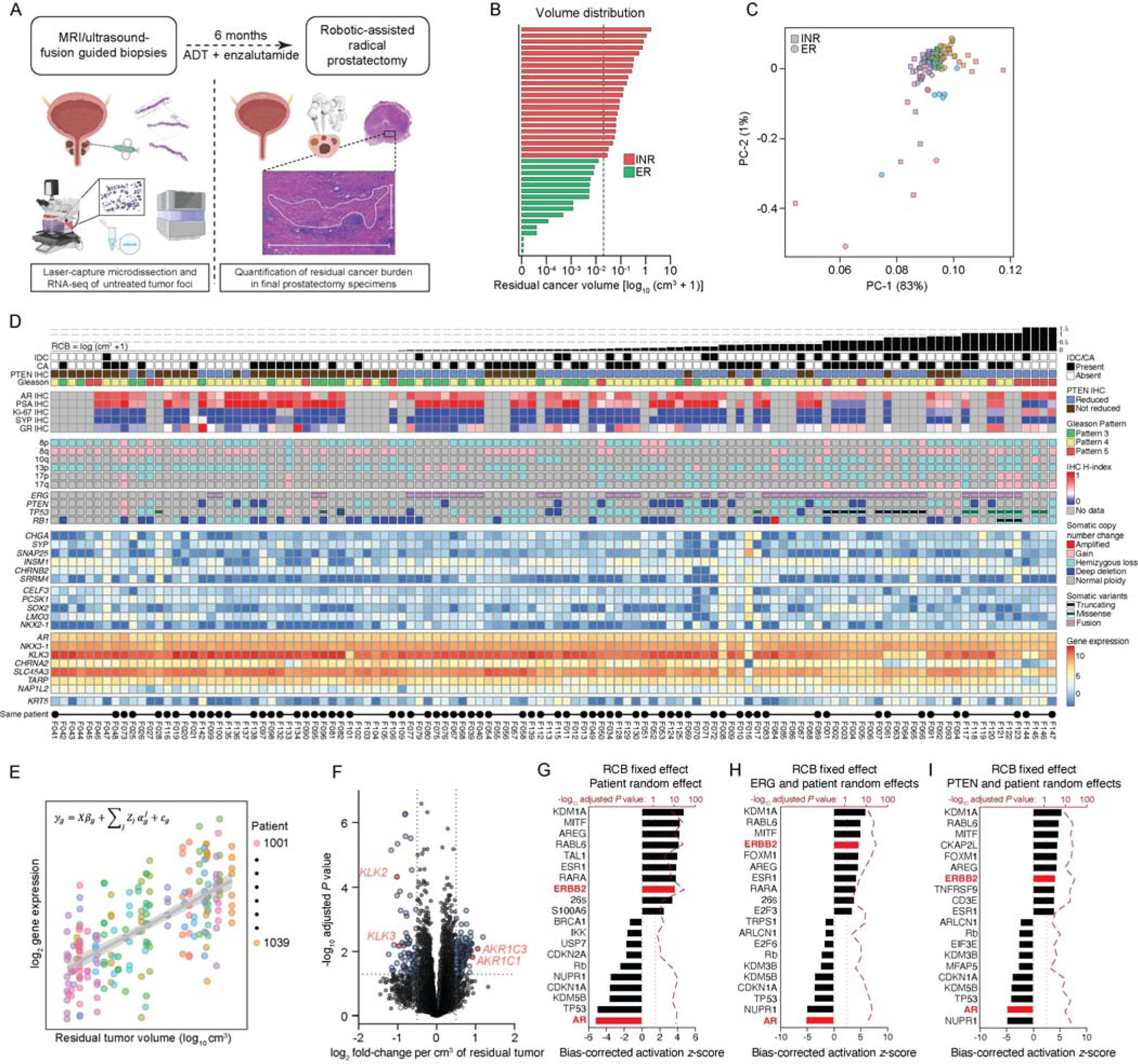
Integrated molecular landscape of prostate tumors prior to neoadjuvant intense androgen deprivation therapy. **(A)** Schematic of workflow in which laser capture microdissection and RNA-seq of tumor foci from image-guided baseline biopsies (left) are used to assess gene expression differences that track with posttreatment pathologic tumor volumes (right). **(B)** Distribution of residual cancer burden (RCB, one row per patient) plotted on a logarithmic *x*-axis with a pseudocount (cm^3^ + 1). Green bars represent exceptional responders (ER) and red bars represent incomplete and nonresponders (INR) who harbored residual tumor volumes greater than 0.05 cm^3^. **(C)** Principal component analysis of 147 baseline tumor foci transcriptomes. Each dot is colored by patient, with squares representing foci from INR patients and circles representing foci from ER patients. **(D)** Heatmap and oncoprint depicting molecular and histologic features of baseline tumors where each column represents one laser capture microdissected tumor focus subjected to whole-transcriptome sequencing. Identical values are given IHC profiling performed on a single tissue that was subdivided for sequencing. Black bars at the bottom indicate multiple samples from the same patient. Samples are ranked from left-to-right by patient-level residual cancer burden (RCB) volumes. **(E)** Linear mixed-effect model depicting variance in gene expression across samples within each patient (by color) vs. RCB (*x-*axis), showing gene expression pattern for positively-correlating genes. **(F)** Volcano plot depicting differentially-expressed genes (DEGs) determined using a linear mixed-effect model using RCB as a fixed effect and each patient as a random effect. Horizontal boundary depicts the *P* = 0.05 (adjusted) cutoff, and vertical boundaries demarcate genes with a fold-change of at least ±2. DEGs are quantified as fold-change per unit of post-treatment tumor volume (in cm^3^) where genes to the right are more expressed at baseline in tumors with higher volumes and genes to the left are less expressed. **(G-I)** All statistically significant DEGs (*P*_adj_ < 0.05) from the linear mixed-effect model were processed with the upstream regulator module of Ingenuity Pathway Analysis. The ten most activated and inactivated pathways (with adjusted *P* values less than 0.05) are shown for DEG analyses in which **(G)** patients were the only random effect, **(H)** patients and ERG status were random effects, and **(I)** patients and PTEN status were random effects. The bias-corrected *z* score is shown on the bottom *x* axis and the adjusted *P* value is shown on the top *x* axis (-log_10_ transformed).

Our overall goal was to identify variability in genes and pathways expressed at baseline that would track with resistance to therapy. Therefore, we initially stratified samples by our predefined cutpoint of posttreatment residual tumor volume of 0.05 cm^3^, which was measured from the largest area of tumor in the final prostatectomy specimens (**Figs. 1A,B**, and see methods) [8]. As we also sequenced foci from resolving MRI lesions that were acquired from nonresponding patients, we further limited our analysis to 117 foci isolated only from MRI index lesions. By principal component analysis, however, there was no obvious structure that distinguished foci from exceptional responder (ER) patients vs. incomplete and nonresponder (INR) patients (**Fig. 1C**). In addition, application of four predefined gene groups that had previously been shown to segregate treatment resistant tumors into transcriptionally defined subtypes in the castration-resistant setting [38] did not prominently identify multiple baseline tumor subgroups (**Fig. 1D**). Rather, nearly every focus showed strong enrichment for genes in the androgen receptor-dependent adenocarcinoma gene group, which included *AR*, *NKX3-1* and *KLK3* (**Fig. 1D**).

Nevertheless, we noted a trend in which cases that harbored greater volumes of residual tumor after therapy (RCB, toward the right of the heatmap in **Fig. 1D**) expressed lower relative levels of genes in the androgen receptor-dependent adenocarcinoma group, including *KLK3*. This was confirmed by a negative correlation between the H-index of IHC against PSA on the same tissues, the protein product of *KLK3*, when compared to residual cancer volumes on a per-patient level (ρ = -0.43, *P* = 0.016) [22]. Therefore, we sought to use RCB as a continuous variable to identify differentially-expressed genes across the cohort. Retaining the variability afforded by multiply-sampled cases, we employed a linear mixed-effect statistical model, holding RCB as a fixed effect, and modeling each patient as a random effect (**Fig. 1E**). The outcome of this analysis identified 644 differentially-expressed genes (*P*_adj_ < 0.05, see **Supplementary Table 1**), where each unit of fold-change represents 1 cm^3^ of posttreatment RCB (**Fig. 1F**). Amongst the most negative fold-change genes (*i.e.* the genes that were more down-regulated in patients who went on to exhibit exceptional responses) were the AR-responsive *KLK2* and *KLK3* suggesting that these tumors had greater AR activity. By contrast, two of the most positive fold-change genes were *AKR1C1* and *AKR1C3* of the testosterone biosynthesis pathway, representing in part an adaptive response to low androgens in more aggressive prostate tumors [39].

To identify targetable pathways that are overrepresented in baseline tumors that resist therapy, we processed the 644-gene list using the Upstream Regulator module of Ingenuity Pathway Analysis. As depicted in **Figure 1G**, the most negatively enriched (inactivated) regulator was AR (*z*_corr_ = -5.2, *p*_adj_ = 2.6 × 10^-15^), while the upstream regulator with the lowest adjusted *P* value was *ERBB2* (human epidermal growth factor receptor 2, HER2; *z*_corr_ = 3.7, *p*_adj_ = 2.6 × 10^-37^). As part of our previous analyses, we found that both the *TMPRSS2-ERG* fusion (measured by either IHC or RNA-seq) and *PTEN* loss (IHC) were independently associated with poor pathologic responses (see **Fig. 1D**) and had substantial effects on global transcription [22, 40]. Therefore, we further modeled ERG or PTEN IHC status as additional random effects in our linear mixed-effect model. Regressing out the transcriptional impacts of ERG (**Fig. 1H**) nor PTEN (**Fig. 1I**) did not appreciably change AR’s position or statistical significance in the bottom 10 inactivated pathways and HER2’s position in the top 10 activated pathways. Thus, HER2 activity represents potential and distinctive mechanism of intrinsic resistance to neoadjuvant intense ADT.

### HER2 protein expression is associated with poor response

HER2 up-regulation or activation has been implicated previously as an adaptive response to androgen deprivation and AR inhibition in metastatic PCa [35, 41]. We therefore performed a series of immunostains against total and phosphorylated (Y1221/1222) HER2 in matched baseline biopsies and posttreatment prostatectomy sections (**Fig. 2A**) to confirm the results suggested by our transcriptional analyses. Across the cohort, we observed variable HER2 and pHER2 expression in both untreated (biopsy; 37 cases, 1–3 slides per case for HER2; 34 cases, 1 slide per case for pHER2) and residual (posttreatment only; 34 cases; 1–8 slides per case) tumor specimens. As expected, the predominant staining pattern observed for total HER2 in both baseline and posttreatment specimens was membranous (**Supplementary Fig. 1**), encompassing both invasive and intraductal tumor foci, although a subset of cases demonstrated predominantly cytosolic staining for HER2.

**Figure 2.**
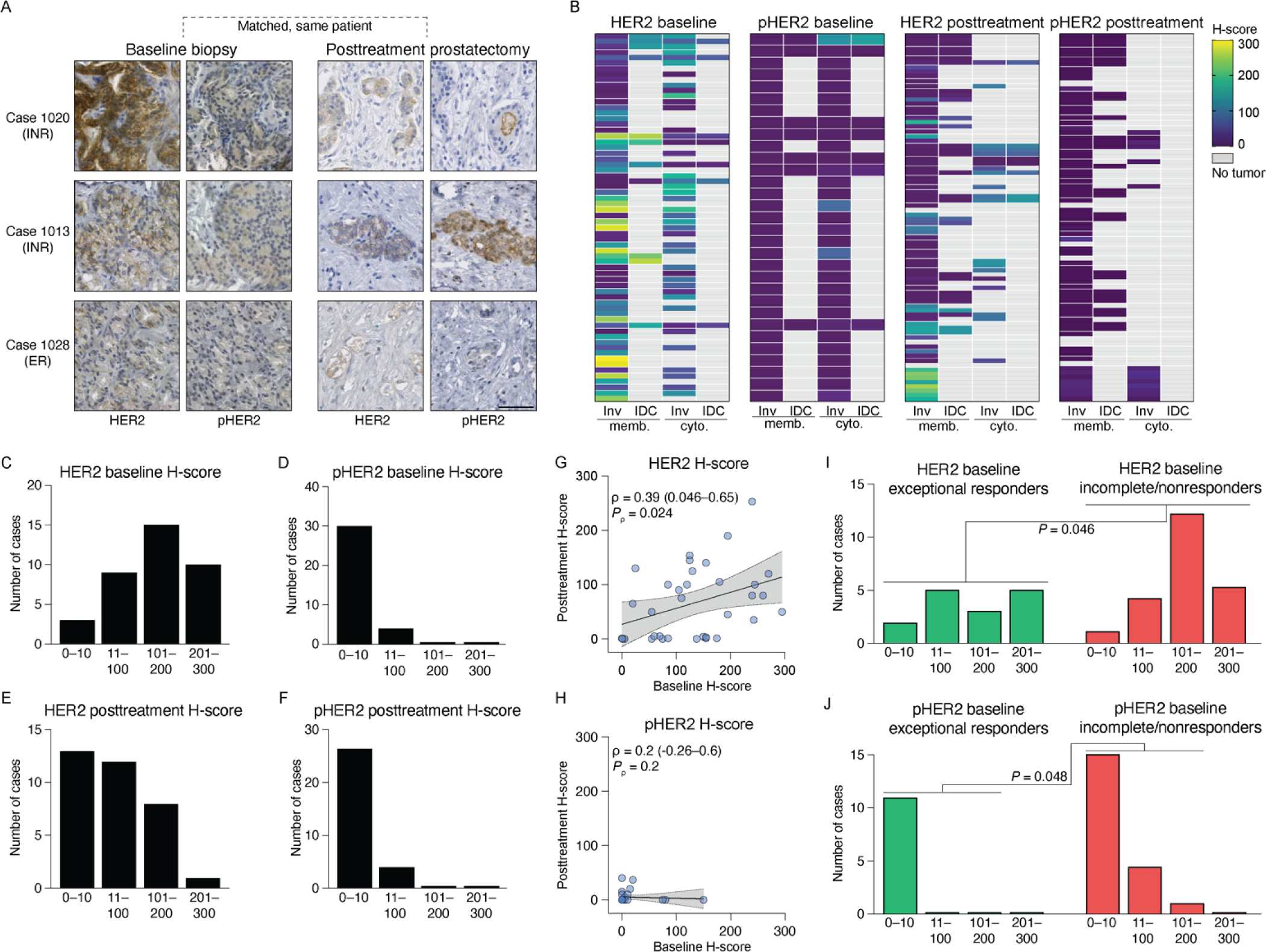
HER2 protein is expressed at baseline in tumor foci that resist therapy and is retained posttreatment. **(A)** Representative micrographs of anti-HER2 and anti-pHER2 IHC in baseline biopsies and residual tumor foci, showing examples from three patients with matched samples. Bar: 50 µm. **(B)** Heatmaps summarizing semi-quantitative analysis of anti-HER2 and anti-pHER2 immunohistochemistry (IHC) performed on entire sections of biopsies and posttreatment surgical specimens. Rows are grouped by patient. **(C-F)** Density plots summarizing the frequency distribution of IHC semi-quantitative analysis, per patient, of baseline biopsies (**C-D)** or of posttreatment prostatectomy specimens **(E-F)** with antibodies against HER2 **(C,E)** and pHER2 **(D,F)**. **(G-H)** Scatter plots showing the association of per-patient HER2 **(G)** or pHER2 **(H)** baseline H-scores (*x*-axis) with posttreatment H-scores (*y*-axis). Statistical significance determined using Spearman’s rank correlation. Line and gray shaded area show the linear regression line and 95% confidence interval for the regression. **(I-J)** Density plots of HER2 **(I)** and pHER2 **(J)** baseline semi-quantitative IHC, stratified by pathologic response in the final surgical specimens with exceptional responders in green and incomplete/nonresponders in red. Statistical significance determined using χ-squared test. Inv: invasive; IDC: intraductal carcinoma; memb: membranous; cyto: cytosolic.

Semi-quantitative H-scoring analysis by an expert genitourinary pathologist (R.T.L.) identified major trends in staining intensity, separately scoring cytosolic and membranous staining for invasive and intraductal tumors, for HER2 and pHER2 immunostains (**Fig. 2B**). Interestingly, HER2 at baseline was expressed mostly in invasive tumor foci (both membranous and cytosolic), and intraductal HER2-expressing baseline tumor were less common that in posttreatment tumors. pHER2 was expressed in a minor fraction of all baseline and posttreatment foci.

We used the maximum H-score for each case, irrespective of morphology, to identify staining trends across the cohort. For example, similar numbers of baseline cases harbored low (H-score 11–100), medium (H-score 101–200) and high (H-score 201–300) intensity for HER2, given that a majority of tumor tissue was positive (**Fig. 2C**). By contrast, most biopsies (30/31) displayed focal pHER2 expression at low and background (H-score 0–10) levels (**Fig. 2D**). Residual tissue (20/34) harbored low and medium intensity for HER2 (**Fig. 2E**) but similar to baseline all tissues expressed pHER2 at low and background levels (**Fig. 2F**).

We observed a statistically significant correlation at the patient-level between pre- and posttreatment IHC staining for HER2 (**Fig. 2G**; ρ = 0.39, *P*_ρ_ = 0.024) but not pHER2 (**Fig. 2H**). Nonetheless, and consistent with our whole-transcriptome analyses, higher semi-quantitative HER2 staining intensities were observed in biopsies from INR vs. ER patients (**Fig. 2I**, *P* = 0.046, χ-squared test). Substantive staining for pHER2 was only observed in biopsies from INR patients (**Fig. 2J**, *P* = 0.048, χ-squared test). Collectively, these data confirm that increased HER2 protein expression at baseline is a molecular feature of tumor that go on to exhibit poor responses to neoadjuvant intense ADT plus enzalutamide, and that this may be associated with canonical HER2 activity.

### Prostate cancer antiandrogen resistance is driven by a pre-existing subpopulation with elevated HER2 activity

Although HER2 activation measured by pHER2 was modestly expressed in tissue samples, we also explored the expression and phosphorylation levels of EGFR and HER3, as persistent or constitutively active signaling through receptor tyrosine kinases occurs in many solid tumor types including subtypes of breast cancer (HER2) and lung cancer (EGFR), especially when the gene is amplified. EGFR expression, but not its phosphorylation (Y1068), displayed similar patterns to HER2 staining (**Supplementary Fig. 2A-E**), although baseline expression stratified by pathologic outcome was not statistically significant (**Supplementary Fig. 2F**, P = 0.10, χ-squared test). HER3 and pHER3 (Y1289) expression was negligible across all samples (**Supplementary Fig. 2G-I**). These findings are consistent with our prior report of exome sequencing of tumor tissue from this study, which did not show any evidence of amplification or mutations rendering it constitutively active for *EGFR* or *ERBB3* (encoding HER3) [22].

We therefore focused on our observation that HER2 expression posttreatment could be due to a subset of resistant cells that were already expressing HER2 prior to the initiation of therapy. Because HER2 expression and its activity tracked with pathologic outcome, we next sought to validate this finding. However, a lack of baseline samples acquired from other studies rendered this infeasible. Nonetheless, our observation that AR activity opposed HER2 activity offered an alternative approach which was amenable to any dataset with whole transcriptome gene expression. For example, when we ranked our original dataset by AR activity using single-sample GSVA of the mSigDB AR Hallmarks gene set, and examined those genes which tracked with AR activity, the Upstream Regulator module of Ingenuity Pathway Analysis reported AR activity to be positively enriched (*z*_corr_ = 5.1, *p*_adj_ = 2.6 × 10^-8^) and *ERBB2* opposing it, now negatively enriched (*z*_corr_ = -2.4, *p*_adj_ = 8.4 × 10^-7^, **Fig. 3A**). We next employed this approach on a novel cohort comprised of 123 tumors samples we acquired from the Prostate Cancer Biorepository Network. We observed similar results (**Fig. 3B**), with AR positively enriched as expected (*z*_corr_ = 5.7, *p*_adj_ = 3.0 × 10^-13^) and *ERBB2* negatively enriched (*z*_corr_ = -3.0, *p*_adj_ = 1.1 × 10^-15^). We also assessed this phenotype in the prostate cancer TCGA cohort, but due to concerns about substantial variability in purity arising from cell type admixtures from the original sample collection [42], we employed deconvolution using dampened weighted least squares (DWLS) estimation to arrive at luminal PCa cell-specific gene expression values. AR was the most activated upstream regular as expected (*z*_corr_ = 6.1, *p*_adj_ = 2.9 × 10^-9^, **Fig. 3C**), and *ERBB2* was again negatively enriched (*z*_corr_ = -2.3, *p*_adj_ = 4.7 × 10^-5^). Collectively, these data suggest that AR activity is inversely associated with HER2 activity across a range of untreated PCa cohorts.

**Figure 3.**
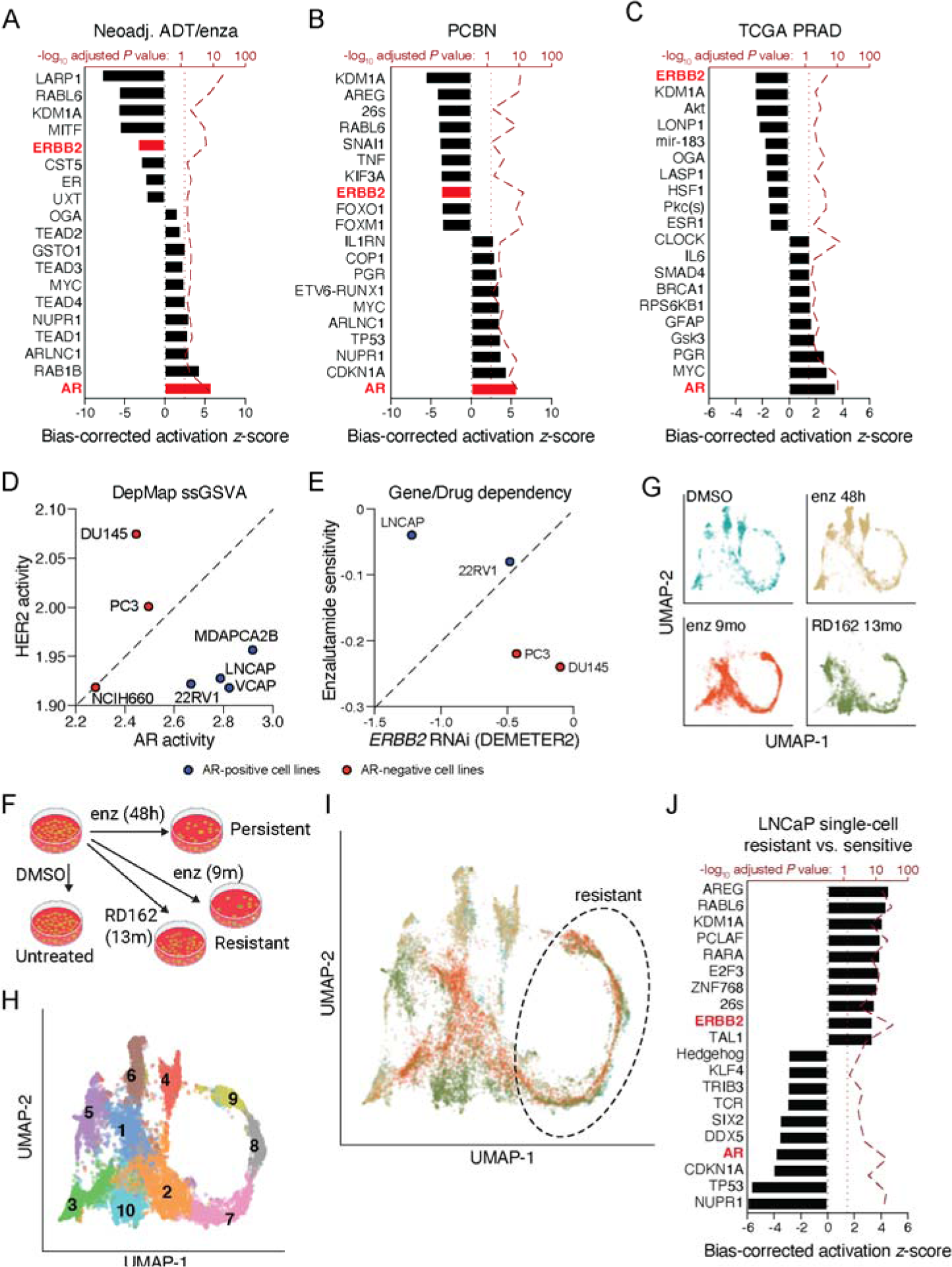
Prostate cancer cell lines recapitulate patterns of enzalutamide resistance observed in patients. **(A-C)** Statistically significant DEGs (*P*_adj_ < 0.05) that were correlated with the “AR Hallmarks” mSigDB geneset processed by single-sample GSVA were analyzed with the upstream regulator module of Ingenuity Pathway Analysis for **(A)** our original neoadjuvant ADT plus enzalutamide cohort, **(B)** 123 tumors from the Prostate Cancer Biorepository Network (PCBN) and the **(C)** prostate cancer TCGA. A linear mixed-effect model was employed with the neoadjuvant cohort in **(A)** modeling repeated measures from patients as random effects. The ten most activated and inactivated pathways (with adjusted *P* values less than 0.05) are shown. **(D-E)** Publicly available data from the Broad DepMap is shown, in which AR-positive cell lines are depicted in blue and AR-negative cell lines depicted in red. Gene expression were summarized using single-sample GSVA for AR and HER2 activity signatures from mSigDB (**D**), and cell death (sensitivity) were plotted to compare matched enzalutamide sensitivity or *ERBB2* RNAi survival scores (**E**). (**F-J**) Publicly accessible single-cell gene expression data from LNCaP cells treated with antiandrogen (**F-G**) were downloaded and normalized together. (**G-I**) UMAP projections of each treatment condition individually (**G**), clustered by differential expression (**H**), and overlaid, colored by treatment condition (**I**). Following trajectory and pseudobulk differential expression analysis, statistically significant DEGs (*P*_adj_ < 0.05) in the “resistant” clusters were analyzed with the upstream regulator module of Ingenuity Pathway Analysis. The ten most activated and inactivated pathways (with adjusted *P* values less than 0.05) are shown.

Using data from the Broad Institute Dependency Map (DepMap), we explored the relationship between AR or HER2 activity (similarly measured using single-sample GSVA), *ERBB2* expression, and enzalutamide sensitivity. AR activity stratified several established cell lines, with NCIH660, PC3 and DU145 cells showing lower signature scores (**Fig. 3D**). Two of these lines, DU145 and PC3 cells, also had greater HER2 scores, while the AR high cell lines (22Rv1, VCaP, LNCaP and MDa-PCa-2b cells) uniformly had lower HER2 scores. When comparing the four cell lines that had paired viability data from drug treatment (enzalutamide) and RNAi exposure (against *ERBB2*), LNCaP and 22Rv1 cells, which are AR-positive, display greater sensitivity to enzalutamide than PC3 and DU145 cells (**Fig. 3E**). However, PC3 and DU145 demonstrated impaired growth (relative to LNCaP) when treated with RNAi against *ERBB2*, indicating that established cell lines may be an acceptable model to explore the AR-HER2 relationship further (**Fig. 3E**).

We accessed a published single-cell RNA-seq dataset [43] in which LNCaP cells were treated with antiandrogen and transcriptomically profiled shortly after exposure (48h) or after resistance developed (9-13 months, **Fig. 3F**). After joint normalization of the untreated (DMSO) control and treated samples, we employed trajectory analysis to identify differentially-expressed clusters of genes (**Fig. 3G,H**), with clusters 7, 8 and 9 best representing cell clusters present at baseline that were enriched upon resistance development (**Fig. 3I**). We derived differentially-expressed genes using pseudo-bulk analysis, comparing these clusters against the rest. Ingenuity Pathway Analysis recapitulated our earlier findings, showing *ERBB2* activity increased in resistant cells (*z*_corr_ = 3.0, *p*_adj_ = 7.4 × 10^-38^) while AR activity decreased in sensitive cells (*z*_corr_ = -3.5, *p*_adj_ = 1.6 × 10^-24^). These findings indicate that *de novo* PCa cell resistance to antiandrogen is driven, at least in part, by a pre-existing subset of cells with elevated HER2 activity.

### AR-positive prostate cancer cell lines are sensitive to HER2 inhibition which enriches for cells with greater AR activity

The increased abundance of HER2 activity in enzalutamide-resistant PCa cells raises the possibility that treatment with inhibitors against HER2 (or other receptor tyrosine kinases, RTKs) may either increase tumor cell AR activity or enrich for the population of tumor cells that is harboring greater AR activity. We thus assessed the sensitivity of 8 different RTK inhibitors (RTKi) against three different AR-positive prostate cancer cell lines: LNCaP, LAPC4 and 22Rv1 cells. LNCaP and LAPC4 represent an earlier state of hormone sensitivity in an untreated tumor, while 22Rv1 cells are AR-positive cells with treatment-acquired resistance to hormone therapy and antiandrogen.

Using a 7-concentration dose response curve in sextuplicate, we derived IC_50_ values for each RTKi over a five-day time course, repeated at least three different times (**Fig 4A,B** and **Supplementary Table 2**), assessing viability using Cell-Titer Glo. As predicted above, all three AR-positive cell lines displayed exquisite sensitivity to neratinib (IC_50_: 0.47–1.3 µM), an irreversible inhibitor selective for HER2. Cells were also sensitive to afatinib (IC_50_: 1.0–2.8 µM), an inhibitor of homo/heterodimerization for EGFR/HER2/HER3. Consistent with our finding that EGFR alone did not track with resistance (see **Fig. 1**), EGFR-specific inhibitors such as gefitinib and erlotinib displayed weaker or no (respectively) antitumor effect (**Fig. 4B**).

**Figure 4.**
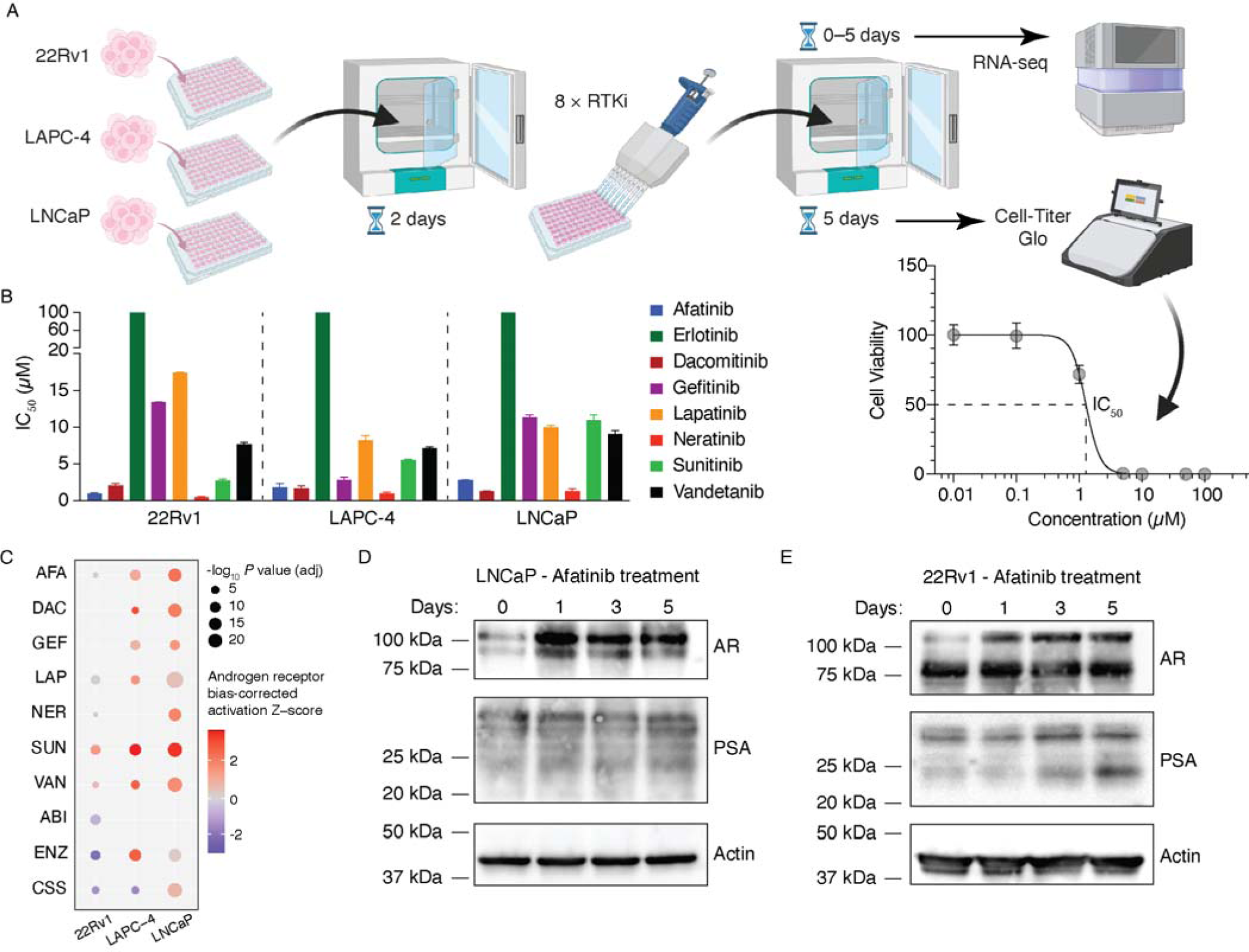
HER2 inhibition selects for prostate cancer cells with greater AR activity. **(A)** Schematic depiction of *in vitro* screening of 22Rv1, LAPC-4 and LNCaP cells with eight different receptor tyrosine kinase inhibitors (RTKi). (**B**) Half-maximal inhibitory concentrations (IC_50_ values) are shown for each RTKi used, per cell line. Values depict the average value derived from at least three dose response curves derived for the individual lots of each RTKi that was used. (**C**) Each cell line was treated with the indicated RTKi, abiraterone (ABI), or enzalutamide (ENZ) at the IC_50_ derived for that lot of drug. Charcoal stripped serum (CSS) was used in place of FBS (see methods) in cell culture media. Treatments were performed over a 5-day time course and samples were acquired days 0, 1, 3 and 5. RNA extracted from each sample (performed in duplicate) was subjected to whole-transcriptome sequencing, with differentially expressed genes (correlating with time) processed using the upstream regulator module of Ingenuity Pathway Analysis. (**D-E**) Western blots depicting protein levels of LNCaP cells (**D**) and 22Rv1 cells (**E**) treated with afatinib (at its empirically determined IC_50_) for 0–5 days. Blots shown are representative of at least three independent experiments. Actin is shown as a loading control.

Using these empirically determined IC_50_ doses, we next examined the impact of drug exposure on gene expression over a 5-day time course across three independent experiments. As controls, we also treated with enzalutamide, abiraterone, or grew cells with charcoal-stripped serum to deplete the media of androgens. We extracted RNA from each time point and performed whole-transcriptome sequencing (**Fig. 4A**). Using a linear mixed-effect model we identified differentially expressed genes that changed in the same direction (up or down) over the entire time course, per unit time. We then processed these genes using the Upstream Regulator module of Ingenuity Pathway Analysis to determine if AR activity was increasing on account of the RTK treatment (**Fig. 4C**). Most RTKi treatments resulted in statistically significant and positive *z* scores for AR activity. Notably, we observed that upon treatment with afatinib at each cell line’s IC_50_, increased AR pathway activity was observed across the LNCaP, LAPC4 and 22Rv1. Increases for AR activity were also observed with neratinib treatment for LNCaP and 22Rv1 cells, and surprisingly, treatment with the VEGFR inhibitor sunitinib also had a consistent and positive impact on AR activity. By contrast, treatment with ADT (CSS), abiraterone or enzalutamide had mostly negative effects on AR activity.

At the protein level, treatment with afatinib in both LNCaP (**Fig. 4D**) and 22Rv1 (**Fig. 4E**) cells enriched for AR expression after one day and persisted for up to five days. Increases in PSA protein levels were also observed over days 1–5 (**Fig. 4D-E**). Collectively, these findings suggest that receptor tyrosine kinase inhibitors, particularly the HER2 inhibitors afatinib and neratinib, are highly potent on established AR-positive PCa cell lines, and that cells resistant to these treatments after 1–5 days express proportionally greater levels of AR.

### Nascent prostate cancer harbors distinct AR-positive and HER2-positive subpopulations

Given the potency of HER2 inhibition in PCa cell lines, we next sought to determine whether HER2 inhibition had an antitumor effect in human patient-derived PCa models. Therefore, we treated four different PCa organoid models derived from intermediate-risk disease with 0–5 µM neratinib or 10 µM ABT-373 (BCL-2 inhibitor) as a positive control. 48 hours after treatment, we dissociated the organoids, stained the cells with 7-aminoactinomycin D (7-AAD) and conjugated antibodies against Annexin V, and quantified staining using flow cytometry. After gating each organoid model for live and dead cells (**Fig. 5A**), we applied these gates to the neratinib-treated models (**Fig. 5B**). As summarized in **Figure 5C**, two models (183 and 187) demonstrated exquisite sensitivity to neratinib, with >80% of cells dead at 3 µM and >95% cells dead at 5 µM. By contrast, model 190 was mostly resistant, with less than 35% apoptotic at 5 µM. Intriguingly, model 188 displayed a mixed phenotype with approximately 50% and 65% of cells apoptotic at 3 µM and 5 µM, respectively. This range of responses suggests that the proportion of HER2-dependent cells may change between individuals, and that pre-existing subpopulations of each tumor may display differential sensitivity.

**Figure 5.**
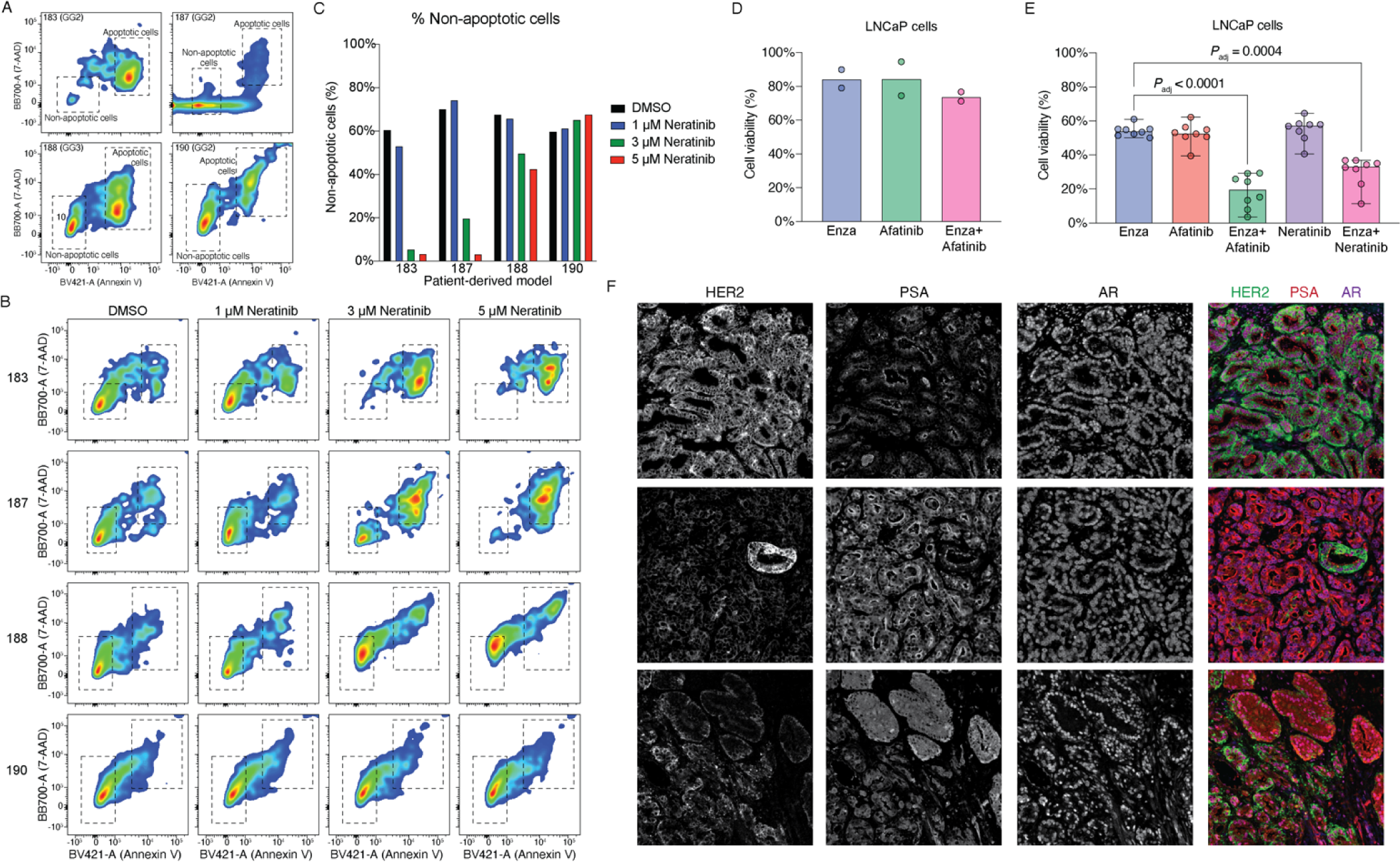
Prostate cancer cells expressing high levels of HER2 are distinct from tumor cells with high levels of AR activity. **(A)** Flow cytometry of four human prostate cancer organoids treated with 10 µM ABT-373 to identify the gates for apoptotic and nonapoptotic cells. The pathologic grade group of each of the organoid models (183, 187, 188 and 190) is also shown. **(B)** Flow cytometry of the same four human prostate cancer organoids as in (A), treated with DMSO, 1 µM neratinib, 3 µM neratinib or 5 µM neratinib for 48 hours. The gates defined in (A) were used to identify the apoptotic and nonapoptotic cells. **(C)** Bar graph showing the proportion of nonapoptotic cells measured in (B). **(D)** LNCaP cells were treated with either enzalutamide or afatinib ± enzalutamide, (at their respective IC_20_) for five days. Cell viability was measured using Cell-Titer Glo. Data shown is the average of two experiments. **(E)** LNCaP cells were treated with enzalutamide, afatinib ± enzalutamide, or neratinib ± enzalutamide for five days. Cell viability was measured using Cell-Titer Glo. Data shown is the median of at least three replicate experiments. Error bars 95% confidence interval for experiments with at least 5 replicates. Statistical significance was measured using a repeated-measures ANOVA test with Bonferroni adjustment for multiple comparisons. **(F)** Multiplex immunofluorescent micrographs (taken at 20 × magnification) of three representative human prostate tumors stained with HER2 (green), PSA (pink) and AR (purple).

We reasoned that if pre-existing and co-mingled cell populations were to be treated using a combination approach, the exposure paradigm would be additive rather than synergistic. Indeed, a combination approach of enzalutamide and afatinib in LNCaP cells at the drugs’ respective IC_20_ marginally increased decreased cell viability greater than the IC_20_ dose of enzalutamide or afatinib alone (**Fig. 5D**). By contrast, treatment with enzalutamide, afatinib or neratinib at an IC_50_ dose consistently killed approximately 50% of LNCaP cells, and the addition of afatinib or neratinib to enzalutamide at its IC_50_ further improved cell killing to approximately 80% (*P*_adj_ < 0.0001, repeated measures ANOVA) and 65% (*P*_adj_ = 0.0004), respectively (**Fig. 5E**). Collectively, these data indicate that the improved cell killing by combining antiandrogen with HER2 inhibition is due to the simultaneous targeting of two distinct cell subpopulations.

Finally, we asked whether such a variable proportion of AR activity-high or HER2-high prostate tumor cells was a universal feature of untreated (hormone-sensitive) localized prostate tumors. We devised a multiplex immunofluorescent panel with antibodies against AR, PSA and HER2 and extended our analysis to an institutional cohort of 84 radical prostatectomy tissues that received no prior androgen deprivation therapy. As shown in the representative micrographs in **Figure 5F**, although PCa luminal epithelial cells were AR-positive, the intensity of PSA and HER2 varied substantially. Importantly, regions that were PSA (and/or AR) high were mutually exclusive to tumor foci that were expressing high levels of HER2, and *vice versa*. These findings indicate that localized prostate cancers may be variably sensitive to HER2 inhibition, or combination AR and HER2 inhibition, depending upon the proportion of HER2-high AR activity-low tumor cells contained therein.

## DISCUSSION

Neoadjuvant therapies for prostate cancer face inherent challenges of patient selection and the uncertainties of how subsequent therapies, upon recurrence, contribute to metastasis development. In the current study, we identified patterns of baseline gene expression signatures that tracked with the volume of posttreatment residual disease, a surrogate pathologic biomarker for BCR [20]. Our identification of HER2 activity association with poor responses to intense androgen deprivation neoadjuvant therapy is in turn associated with these tumors’ lower dependence on AR and thus decreased sensitivity to AR antagonism. We also demonstrated that HER2 inhibition, alone or in conjunction with AR-targeted therapies, has potent anti-tumor activity.

Significantly, our present study demonstrates that HER2 expression in primary PCa represents a therapeutic vulnerability. HER2 has long been a tantalizing target in PCa, although most of these studies have been conducted in the metastatic and castration-resistant setting after tumors have recurred after androgen ablation [32, 34–37, 44, 45]. One possibility for these studies’ outcomes is that the molecular mechanisms suppressing AR activity in HER2-high tumor cells become sufficiently reprogrammed after years of prolonged ADT for recurrent PCa such that the therapeutic window for co-targeting HER2 and AR has been lost. Indeed, the *AR* gene is amplified in 50-80% of ADT-treated metastatic tumors, which results in significant reprogramming of cistromic and *cis*-regulatory elements governing gene expression [46, 47]. In the hormone sensitive setting, HER2 monotherapies have also been largely ineffective [44, 48], which may be explained in part by our finding that the proportion of AR- and HER2-dependent cells varies by patient, necessitating co-targeting. Our finding that approximately 60% of localized high-risk patients in our current study harbored moderate-to-high levels of HER2 protein expression in the absence of genomic amplification mirrors those of other histologic analyses [49–51].

Although AR-targeted therapies alone are highly effective in the hormone-sensitive localized and metastatic settings, the addition of chemotherapy to ADT plus a novel hormonal agent has demonstrated that up-front intensification delays progression to castration resistance [52, 53]. With genomically-informed neoadjuvant studies (*e.g.* NCT04812366) testing similar combinations in the presurgical setting, our current report substantiates clinical investigation into the combination of HER2 with or without intensive ADT, based in part on AR high *vs.* low transcriptional activity or the proportion of each cell subset as determined by IHC. Indeed, this proportion may serve as a molecular determinant of ADT sensitivity, given that the AR activity-low cells are a pre-existing subpopulation necessitating combination therapy at doses indicative of two independent targets (see **Figs. 4 and 5**). Thus, as this subset of never-before-treated, *de novo* prostate tumors are showing little (or no) AR activity by transcriptional and histologic profiling, ADT may not be appropriate as a universal first-line systemic therapy.

A distinction, as well as a limitation, of the present study was our use of prostate biopsy tissue from a neoadjuvant therapy clinical trial as a platform for discovery. Such abundance of biopsy tissue, made possible by advances in targeted biopsy technologies and the requirement of our treatment protocol for on-study biopsies, was not possible for other neoadjuvant studies with molecular components recently published [10, 11, 24, 25]. However, without well-defined transcriptomes across a broad range of tumor pathologic responses (RCBs), direct validation of the results of this study was not possible in a statistically meaningful secondary cohort. However, by using AR transcriptional activity as a surrogate marker of RCB, we found that AR activity and HER2 activity remain as distinctly inverse properties of prostate tumors which could be extended to other case sets. This inverse relationship of AR and HER2 activity, which we describe here in localized prostate cancer for the first time, is similar to that recently reported for salivary gland cancers [54, 55]. Salivary duct carcinoma (SDC) normally expresses the androgen receptor, but a subset was shown to have higher levels of HER2 which corresponded to low or absent AR expression [55]. Consistent with the current study, patients with high HER2/low AR SDC had worse prognoses [55].

Determining long-term disease-free survival benefits requires more extensive follow-up from the results of neoadjuvant therapy clinical trials, especially when conducted as platform studies with pathologic readouts. However, as we have shown here, a feature of prostate cancer cells to resist direct AR antagonism by enzalutamide is their intrinsic ability to have lower levels of AR activity. Therefore, even in the absence of extended clinical follow-up data, we have identified novel features of prostate tumor cells with direct clinical significance in their sensitivity to HER2 inhibition while demonstrating resistance to enzalutamide. Going beyond the localized high-risk population, we propose that the AR-low population of metastatic hormone-sensitive PCa that would otherwise show poor response to enzalutamide may be targeted by HER2 inhibition, a hypothesis that could be readily tested in a genomically-guided neoadjuvant therapy clinical study.

## PATIENTS, MATERIALS AND METHODS

### Ethical approval

Human biospecimens used in this study were derived from tissues acquired as part of National Cancer Institute study 15-c-0124 (NCT02430480), which was approved by the National Institutes of Health Institutional Review Board (IRB) in accordance with the Declaration of Helsinki. Tissue samples were acquired from the Prostate Cancer Biorepository Network (PCBN) through an agreement with the University of Washington Genitourinary Cancer Specimen Biorepository (agreement number 888). Prostate cancer patient-derived organoids were acquired and grown at Emory University under an Emory IRB-approved protocol (STUDY00005649). Standard-of-care prostatectomy specimens were used in accordance with protocols 15-008 and 15-492 from the Dana-Farber/Harvard Cancer Center.

### Biopsy and radical prostatectomy selection

Biopsy specimens for laser capture microdissection (LCM) and immunohistochemistry (IHC) were previously described [22]. Briefly, patients who enrolled in a clinical trial of neoadjuvant ADT plus enzalutamide for 6 months underwent templated and MR/US-fusion targeted biopsies prior to the initiation of therapy. Biopsies were selected for LCM and IHC based on tumor content and adverse pathologic features. Radical prostatectomy specimens from each patient were used to derive residual cancer burden (RCB) volumes as a measure of absolute pathologic response, as previously described [8, 22]. Briefly, RCB was calculated by multiplying the number of slices containing residual tumor by the largest cross-sectional width and length of that tissue and by block thickness (0.6 cm). Volume was further corrected by multiplying by 0.4 to account for tumor cellularity. We considered RCB < 0.05 cm^3^ as exceptional responders (ER) and we grouped incomplete and nonresponders (INR) into a single category for RCB > 0.05 cm^3^. Residual tumor was identified using a combination of routine H&E additional immunostains to verify the presence of residual tumor, including anti-NKX 3.1, PIN-4 cocktail, CAM5.2, and anti-p63.

All prostatectomy blocks harboring residual tumor were used for immunostains against EGFR, pEGFR, HER2, pHER2, HER3 and pHER3. One biopsy block per case was used for immunostains against HER2. Cases with biopsy blocks harboring sufficient remaining tumor material for additional IHC: anti-EGFR: 35; anti-pHER2: 31; and anti-pEGFR: 29.

A separate cohort comprised of radical prostatectomy specimens from 84 patients (described previously in [56]) receiving no prior therapy was used for multiplex staining of HER2, AR and PSA. Each block containing tumor tissue was selected from the index lesion of each tumor.

Prostate cancer patient-derived organoids were acquired from fresh prostatectomy tissue in patients undergoing radical retropubic surgery at Emory St. Joseph Hospital (Atlanta, GA). Tumor tissue was identified jointly by the attending surgeon and pathologist at the time of tissue grossing.

Frozen prostatectomy specimens (embedded in OCT) from the PCBN were sectioned onto glass slides and stained with H&E, or were shipped as ribbon curls in microfuge tubes. After review by a genitourinary pathologists (R.T.L.) to confirm the presence of tumor cells, ribbon curls were processed using the RNeasy Plus Mini Kit (Qiagen) to extract total RNA.

### Tissue processing and immunohistochemistry

Diagnostic H&E slides of biopsies fixed in formalin and paraffin-embedded (FFPE) were reviewed by two genitourinary pathologists (R.T.L. and H.Y.). Additional immunostains for tissue characterization and/or to guide laser capture microdissection (LCM) were performed on 5-µm serial sections of tissue as previously described [22]. All IHC assays were performed using validated protocols on an PATH FLX autostainer (Biocare Medical). For LCM, additional 5-µm serial sections of tissue were cut onto polyethylene naphthalate membrane slides (MicroDissect GmbH), with glass slides cut after every five membrane slides for additional H&E and IHC stains to serve as references. Membrane slides were briefly baked, deparaffinized, rehydrated and stained with Paradise stain (ThermoFisher) according to the manufacturer’s protocol. Approximately 10,000-50,000 tumor cells per ROI were captured from serial sections using an ArcturusXT Ti microscope onto CapSure Macro LCM Caps (Thermo Fisher) using the infrared capture and ultraviolet cutting lasers. Adjacent stromal tissue that was incidentally captured was ablated using the UV laser. Micrographs of each cap were taken after each LCM session and cross-referenced against reference slides by two blinded genitourinary pathologists (R.T.L. and H.Y.) to verify the regions captured. RNA was extracted from LCM tissues by using a clean scalpel to excise the LCM cap polymer and immerse it in Buffer PKD from the RNeasy FFPE Mini Kit (Qiagen).

For IHC against EGFR, pEGFR, HER2, pHER2, HER3 and pHER3, glass slides containing tissue sections were baked for 30 minutes at 60°C. Following deparaffinization in xylenes and rehydration through graded alcohols, antigen retrieval was performed using a NxGen Decloaker (Biocare Medical) at 110°C for 15 minutes in Tris-EDTA Buffer (Abcam; ab93684), pH 9.0. Next, a thin border was drawn around the edges of each glass slide using a PAP pen. After 10-minute incubations in Background Punisher (Biocare; BP974), 300 μL of primary antibody solutions were prepared and incubated with tissues at room temperature at 1:100 dilutions into Renoir Red diluent (Biocare Medical; PD904) for 1 h: anti-EGFR clone D38B1 (Cell Signaling; 4267), anti-pEGFR clone D7A5 (Cell Signaling; 3777), anti-HER2 clone 29D8 (Cell Signaling; 2165), anti-pHER2 clone 6B12 (Cell Signaling; 2243), anti-HER3 clone D22C5 (Cell Signaling; 12708), and anti-pHER3 clone 21D3 (Cell Signaling; 4791). Secondary detection was achieved with Mach 4 (Biocare Medical; M4U534H) polymer and/or probe for 30 minutes. Chromogen development was achieved with Betazoid DAB (Biocare Medical; BDB2004) and counterstained with CAT hematoxylin (Biocare Medical; CATHE) diluted 1:2 into distilled water. Slides were dehydrated through graded alcohols into xylene, mounted using Permount (Thermo Fisher), and digitized using a Carl Zeiss AxioScan.Z1 microscope slide scanner equipped with a Plan-Apochromat 20× NA 0.8 objective.

For multiplex immunofluorescence of AR, PSA and HER2, slides underwent preprocessing, antigen retrieval, and blocking as described above. Anti-HER2 clone 29D8 (Cell Signaling; 2165) was diluted 1:100 in 1× Antibody Diluent/Block (Akoya Biosciences; ARD1001EA) before incubating for 1 h. Slides were washed with TBST then incubated with ImmPRESS HRP Goat-anti-Rabbit IgG Polymer Reagent (Vector Laboratories; 30125) for 1 h. Slides were washed with TBST then incubated with Opal 650 (Akoya Biosciences; OP-001005) diluted 1:150 in 1× Plus Amplification Diluent (Akoya Biosciences; FP1498) for 1h. The next day, slides were removed and washed in diH_2_O. Antibodies were stripped using HIER with Diva Decloaker (Biocare Medical; DV2004MX) in a NxGen Decloaking Chamber at 110°C for 30 m. After cooling, slides were loaded onto the PATH FLX autostainer. Slides were washed with TBST, then blocked using Background Punisher for 1 h. AR clone D6F1T (Cell Signaling; 5153) was diluted 1:100 in 1× Antibody Diluent/Block (Akoya Biosciences; ARD1001EA) before incubating for 1 h. Slides were washed with TBST then incubated with ImmPRESS HRP Goat-anti-Rabbit IgG Polymer Reagent (Vector Laboratories; 30125) for 1 h. Slides were washed with TBST then incubated with Opal 520 (Akoya Biosciences; OP-001001) diluted 1:150 in 1× Plus Amplification Diluent (Akoya Biosciences; FP1498) for 1 h. The next day, slides were removed and washed in diH_2_O. Antibodies were stripped using HIER with Diva Decloaker in a NxGen Decloaking Chamber at 110°C for 30 m. After cooling, slides were loaded onto the PATH FLX autostainer. Slides were washed with TBST, then blocked again using Background Punisher for 1 h. PSA clone D6B1 (Cell Signaling; 5365) was diluted 1:50 in Renoir Red Diluent and incubated with tissues for 1 h. After washing with TBST, goat anti-Rabbit IgG (H+L) Secondary Antibody, AlexaFluor 555 (ThermoFisher Scientific; A-21428) was diluted 1:50 in Renoir Red Diluent and incubated for 1 h. Slides were washed with TBST, then removed from the autostainer. Slides were washed with diH2O, then incubated with Vector TrueVIEW Autofluorescence Quenching Kid (Vector Laboratories; SP-8400) for 2 m. Slides were washed with diH_2_O, then incubated with 350 nM DAPI (4′,6-Diamidino-2-Phenylindole, Dihydrochloride) (ThermoFisher Scientific; D1306) for 10 m. Slides were washed with diH_2_O then mounted using ProLong Glass Antifade Mountant (Invitrogen; P36980). As controls for efficient and complete antibody stripping, additional control slides were processed with each one of the primary antibodies described, completing all other steps but omitting the other two secondary antibodies. Slides were then digitized on Carl Zeiss AxioScan.Z1 microscope slide scanner equipped with a Plan-Apochromat 20× NA 0.8 objective equipped with a Colibri 7 flexible light source, and post-processed using ZenBlue (Zeiss).

Fully quantitative IHC analyses against AR, PSA, SYP, GR and Ki-67 were generated using Definiens software and reported previously [22]. Semi-quantitative IHC analyses against EGFR, p-EGFR, HER2, p-HER2, HER3 and p-HER3 were performed using the H-scoring approach, which considers the proportion of tumor cells per slide that display no (0), low (1), medium (2) or high (3) staining intensity. The proportion of cells is multiplied by the intensity factor, for a maximum possible score per slide of 300 (100% of cells at score 3). H-scoring was conducted separately for cytosolic and membranous staining with distinct scores recorded for invasive vs. intraductal tumor histologies. The maximum H-score for each case was used for further comparisons.

### Cell culture and immunoblotting

LNCaP and 22Rv1 cells were purchased from the American Type Culture Collection (ATCC). LAPC-4 cells were a kind gift from Dr. Charles Sawyers. Cells were authenticated by STR profiling (Laragen) every six months. LNCaP and 22Rv1 cells were maintained in RPMI-1640 supplemented with 10% fetal bovine serum, 1% penicillin-streptomycin solution, and 1% L-glutamine. LAPC-4 cells were maintained in IMDM supplemented with 10% fetal bovine serum, 1% penicillin-streptomycin solution, and 1% L-glutamine.

Dose response curves were performed with 22Rv1, LAPC-4 and LNCaP cells for each new lot of drug acquired. Abiraterone, enzalutamide, afatinib, erlotinib, dacomitinib, gefitinib, lapatinib, neratinib, sunitinib, and vendetanib were acquired from the National Cancer Institute Developmental Therapeutics Program Open Chemicals Repository Collection. For each assay, cells were seeded in solid white flat-bottom tissue culture-treated 96-well plates, at 5000 cells per well in a volume of 100 µl. Blank wells were filled with 150 µl of complete media. Unused wells were filled with 75 µl of PBS. Each agent was prepared in DMSO (vehicle) in 50 µl at 3× its intended concentration for a final volume per well of 150 µl. Drug concentrations were tested at 100, 50, 10, 5, 1, 0.1 and 0 µM one day after plating, and cell viability was measured after 5 days of treatment using CellTiter-Glo (CTG) 2.0 Cell Viability Assay Reagent (Promega; G9243). Prior to measurement, plates were left to equilibrate at room temperature for 30 m, 75 µl of CTG reagent was added per well, and read using a Tecan Infinite M200 Pro with 1.5 mm of orbital shaking for 2 m, 10 m of incubation, and luminescence reading with integration time for 1000 ms.

Cells were treated with each agent at its determined IC_50_ (or IC_20_), alone or in combination to measure tumor cell viability, RNA levels, or protein levels. For drug treatments in assessing viability, cells were seeded as described above for dose-response curves, treated one day after plating, and grown for five days prior to incubation with CTG. Representative IC_50_ values from a set of experiments are depicted in **Fig. 4B**. Viability measurements were performed at least 5 times independently. For RNA and protein extraction, cells were seeded in six-well plates at 50,000 cells per well in a volume of 2 ml. Cells were grown to 75% confluency prior to the initiation of drug treatment. Cells were grown for a total of 5 days, treated for 5, 3, 1, or 0 days, in duplicate for RNA and protein extraction, and were performed at least three times independently. RNA was extracted using the RNeasy Plus Mini Kit (Qiagen) following the manufacturer’s protocol, scraping cells directly into Buffer RLT Plus. Protein was extracted by scraping cells into RIPA buffer (Pierce; 89900) supplemented with Halt Protease and Phosphatase Inhibitor Cocktail (Thermo Scientific; 78440). Lysates were incubated on ice for 10 m, vortexed, and centrifuged at 21,000 × *g* for 10 minutes at 4°C. Supernatants were stored at -80°C.

Protein lysates were separated by SDS-PAGE on 4–15% Criterion TGX protein polyacrylamide gels (Bio-Rad) and transferred to nitrocellulose membranes via semi-dry transfer. After 1 h blocking in 10% nonfat dry milk in TBST, membranes were incubated with the following primary antibodies overnight at 4°C, all diluted into TBST with 5% BSA: anti-AR clone D6F11, anti-PSA clone D6B1, and anti-actin clone C4 (Millipore; MAB1501). Membranes were washed and incubated with HRP-conjugated secondary antibodies (1:5000–10000 dilutions) for 1 h, reacted with Clarity Western ECL substrate (Bio-Rad; 1705061) and visualized using the ChemiDoc Touch Imaging System (Bio-Rad).

### Patient-derived organoids

Fresh tumor tissues from prostatectomies were minced and digested in basal medium (Advanced DMEM/F12 supplemented with Glutamax, HEPES and antibiotics) containing 5 mg/mL collagenase type II, and 10 µM Rho-kinase inhibitor (Y-27632) overnight at 37°C. Erythrocytes were lysed in 1 ml of red blood cell lysis buffer for 5 minutes at room temperature, followed by centrifugation at 300 × *g*. The cell pellet was resuspended in growth factor-reduced Matrigel (Corning, 354230) and seeded as ∼20,000 cells in a 40-μl drop in the middle of a 24-well plate. Prostate organoids were cultured in basal medium containing: 50× B-27, 10% R-spondin-conditioned medium, 5 ng/mL EGF, 10 ng/mL FGF-10, 5 ng/mL FGF-2, 1.25 mM N-acetylcysteine, 10 µM Y-27632, 500 nM A-8301, 10 mM nicotinamide, 100 ng/mL Noggin, 1 µM prostaglandin E2, 10 µM SB202190, and 1 nM dihydrotestosterone. Medium was changed every 2-3 days. Organoids appear within 7 days after plating and passaging at a 1:4 dilution every 1–3 weeks with TrypLE Express containing 10 µM Y-27632 followed by mechanical dissociation to single cells.

Organoids were plated at a density of 20,000–30,000 cells per well and treated with ABT-373 (10 µM) and neratinib at 1, 3, and 5 µM for 72 hours. Cells were harvested, spun down, and washed twice with 1× cold PBS. Cells were subjected to viability staining with the Pacific Blue Annexin V Apoptosis Detection Kit with 7-AAD (BioLegend; 640926) following the manufacturer’s recommendations. Samples were acquired on BD FACSymphony A3 instrument and analyzed using FlowJo (version 10) software.

### Library preparation, sequencing, bioinformatic processing and gene expression profiling

Up to 100 ng of RNA extracted from FFPE tissues, up to 1 µg of RNA extracted from frozen tissue sections, and 1 µg of RNA from cell lines were used for preparing paired-end Illumina-compatible RNA-seq libraries. Tissue-derived RNA was first processed using the NEBNext Globin & rRNA Depletion Kit (New England Biolabs), while cell line-derived RNA was processed using polyA capture. Enriched RNA was assembled using the NEBNext Ultra II Directional RNA Library Prep Kit for strand-specific sequencing. All libraries were sequenced on Illumina NovaSeq 6000 S4 flowcells.

Paired FASTQ files were processed as previously described [40]. Briefly, reads were trimmed using Trimmomatic version 0.36, and gene-level counts were estimated using RSEM version 1.3.2 as a wrapper around STAR version 2.7.0f in stranded mode. Gene fusions were identified using defuse version 0.8.1. FASTQ pairs of TCGA data were downloaded from the NCI Genomic Data Commons and processed using an identical pipeline. Raw counts were normalized using the trimmed mean method in edgeR package in R, and used to generate heatmaps with the pheatmap package.

Differentially-expressed genes per unit of residual cancer burden (RCB) were derived using the variancePartition package, modeling units of RCB as the fixed effect, using the formula ∼RCB + (1|Patient) with patient as the only random effect, or ∼RCB + ERG/PTEN (1|Patient) incorporating ERG or PTEN as additional random effects in the linear mixed effect model. Alternatively, AR enrichment score for each sample was calculated using the gene set “Hallmark of androgen response” and with the GSVA package in ssgsea mode with tau = 0.75. Next, differentially-expressed genes (DEGs) associated with AR activity were derived using ∼AR + (1|Patient) in variancePartition. For the TCGA and PCBN cohorts, DEGs were identified using linear regression models with the DESeq2 package.

Gene expression (v.21Q1), drug sensitivity (v.19Q4) and RNAi (Broad/Novartis/Marcotte) viability screen data were downloaded from the Broad Institute DepMap (depmap.org). Gene expression data was processed using the ssGSEA module of GenePattern (genepattern.org) to measure HER2 and AR activity gene sets from mSigDB.

LNCaP single-cell gene expression data was downloaded from the NIH Gene Expression Omnibus using accession ID GSE168668. Data was processed using cellranger version 7.2.0 and aligned to GRCh38-2020-A. The resulting cells were filtered based on total gene counts, number of unique reads and mitochondrial fractions. Next, the data were normalized using computeSumFactors from the scran package and integrated using fastMNN from the batchelor backage. Clustering was performed using Louvain method with clusterCells from the scran package. *K* was calculated using clusGap from the cluster package. Trajectory analysis was performed on the batch-corrected embedding using the slingshot package.

Differential gene analysis from treated prostate cancer cell lines over a time course was performed with formula “∼ day” in voomWithWeights and dream from the variancePartition package. The final bias-corrected *Z* scores were derived by averaging the two replicates from each treatment condition (cell line, drug and day).

### Statistical analysis

Statistical analyses were performed using GraphPad Prism version 10, Microsoft Excel for Mac version 16, and R version 4.2.0. Comparisons of single factors based on residual cancer burden were performed using linear mixed effect models with up to two random effects. Pathway enrichment was filtered and sorted by adjusted *P* values and bias-corrected *Z* scores. Associations between factors were measured using Spearman correlations. Comparison of cell viabilities was performed using ANOVA tests with Bonferroni adjustment for multiple comparisons.

## Data Availability

All data produced are available online at the in the Database of Genotypes and Phenotypes (dbGaP) or Gene Expression Omnibus (GEO) at https://www.ncbi.nlm.nih.gov/gap/ and https://www.ncbi.nlm.nih.gov/geo/, respectively, and can be accessed with phs001938.v3.p1 (dbGaP), phs001813.v3.p1 (dbGaP), GSE183019 (GEO), GSE183100 (GEO), GSE201284 (GEO), GSE183126 (GEO), GSE152516 (GEO), and GSE222196 (GEO).

https://www.ncbi.nlm.nih.gov/geo/

## DECLARATIONS PAGE

### Ethics approval and consent to participate

The collection and analysis of tissue from patients with localized prostate cancer treated by neoadjuvant androgen deprivation therapy plus enzalutamide was approved by the Institutional Review Board of the National Institutes of Health (protocol 15-c-0124, NCT02430480). All patients provided informed consent before participating. This research was conducted in accordance with the principles of the Declaration of Helsinki. Tissue samples were acquired from the Prostate Cancer Biorepository Network (PCBN) through an agreement with the University of Washington Genitourinary Cancer Specimen Biorepository (agreement number 888). Prostate cancer patient-derived organoids were acquired and grown at Emory University under an Emory IRB-approved protocol (STUDY00005649). Standard-of-care prostatectomy specimens were used in accordance with protocols 15-008 and 15-492 from the Dana-Farber/Harvard Cancer Center.

### Availability of data and materials

The data underlying this article have been deposited in Database of Genotypes and Phenotypes (dbGaP) and Gene Expression Omnibus (GEO) at https://www.ncbi.nlm.nih.gov/gap/ and https://www.ncbi.nlm.nih.gov/geo/, respectively, and can be accessed with phs001938.v3.p1 (dbGaP), phs001813.v3.p1 (dbGaP), GSE183019 (GEO), GSE183100 (GEO), GSE201284 (GEO), GSE183126 (GEO), GSE152516 (GEO), and GSE222196 (GEO).

### Authors’ contributions

Data acquisition: S.W., I.M.K., D.L., S.Y.T., J.R.B., N.T.T., A.B., B.V., N.C.W., N.V.C., R.A., R.L.

Methodology: S.W., A.T.K., S.Y.T., J.R.B., A.B., J.M.F., C.S.J.

Reagents: H.T.K., P.A.P., P.L.C., B.T., W.L.D., F.K.

Analysis: S.W., A.T.K., R.T.L., N.T.T., A.B., C.L., H.Y., S.A.H.

Manuscript: All authors Supervision: S.W., H.T.K., A.G.S.

Accessed and verified all underlying data: A.G.S.

## Acknowledgments

The authors gratefully acknowledge the patients and the families of patients who contributed to this study. Portions of this work utilized the computational resources of the NIH HPC Biowulf cluster.

## SUPPLEMENTARY FIGURES

**Supplementary Figure 1.**
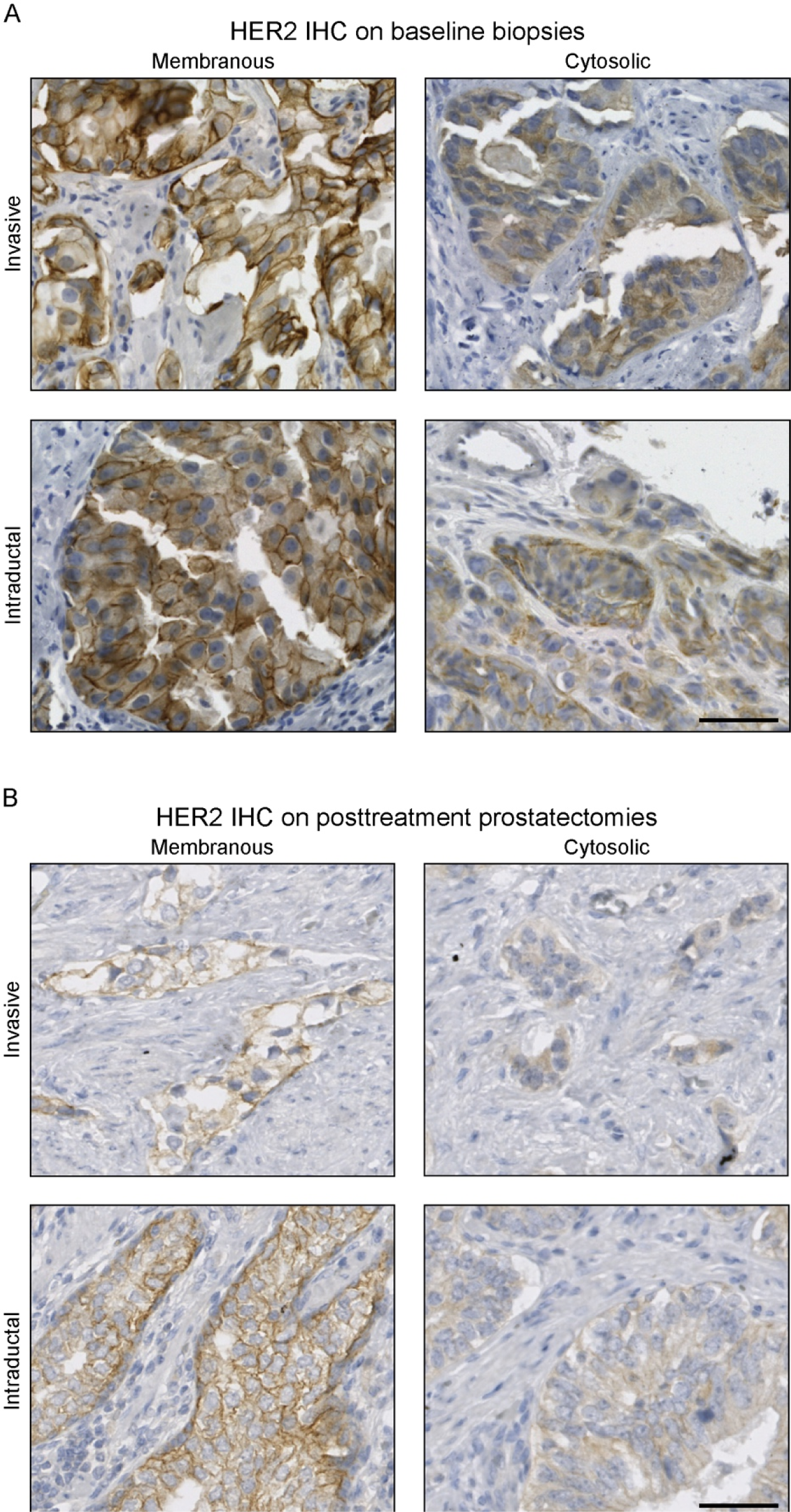
Representative micrographs of anti-HER2 IHC. Examples are shown of invasive and intraductal morphologies with either membranous or cytosolic staining in baseline biopsy **(A)** and posttreatment radical prostatectomy **(B)** specimens. Bar: 50 µm.

**Supplementary Figure 2.**
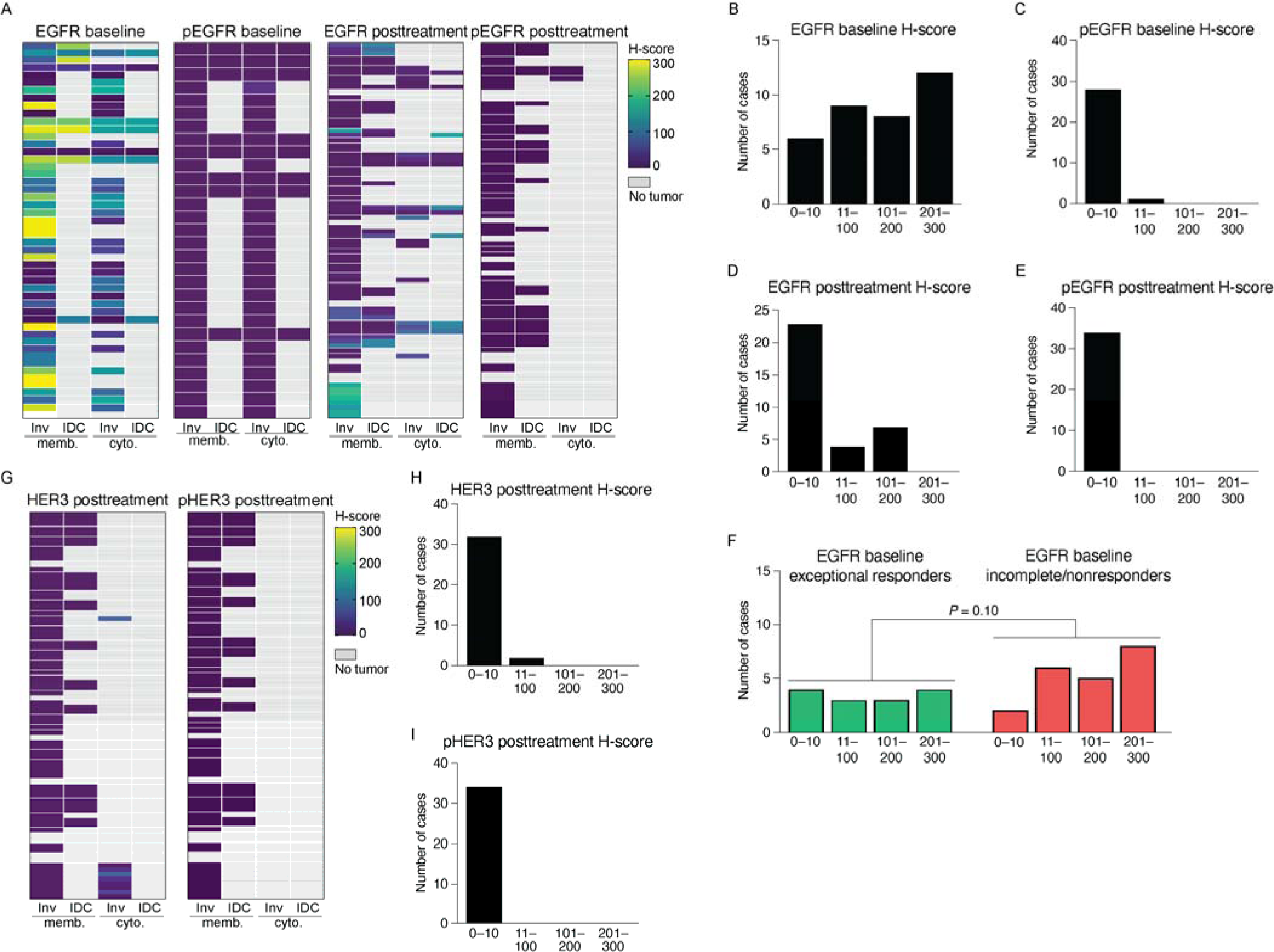
Expression and phosphorylation of EGFR and HER3 are not associated with pathologic response. **(A)** Heatmaps summarizing semi-quantitative analysis of anti-EGFR and anti-pEGFR IHC performed on entire sections of biopsies and posttreatment surgical specimens. Rows are grouped by patient. **(B-E)** Density plots summarizing the frequency distribution of IHC semi-quantitative analysis, per patient, of baseline biopsies (**B-C)** or of posttreatment prostatectomy specimens **(D-E)** with antibodies against EGFR **(B,D)** and pEGFR **(C,E)**. **(F)** Density plots of EGFR baseline semi-quantitative IHC, stratified by pathologic response in the final surgical specimens with exceptional responders in green and incomplete/nonresponders in red. Statistical significance determined using χ-squared test. **(G)** Heatmaps summarizing semi-quantitative analysis of anti-HER3 and anti-pHER3 IHC performed on entire sections of posttreatment surgical specimens. **(H-I)** Density plots summarizing the frequency distribution of IHC semi-quantitative analysis, per patient, of posttreatment tumors with antibodies against HER3 **(H)** and pHER3 **(I)**. Inv: invasive; IDC: intraductal carcinoma; memb: membranous; cyto: cytosolic.

## REFERENCES

1. Haffner, M.C., et al., Genomic and phenotypic heterogeneity in prostate cancer. Nat Rev Urol, 2020.

2. Ye, H. and A.G. Sowalsky, Molecular correlates of intermediate- and high-risk localized prostate cancer. Urol Oncol, 2018. 36(8): p. 368–374.

3. Moris, L., et al., Benefits and Risks of Primary Treatments for High-risk Localized and Locally Advanced Prostate Cancer: An International Multidisciplinary Systematic Review. Eur Urol, 2020. 77(5): p. 614–627.

4. Jones, C.U., et al., Radiotherapy and short-term androgen deprivation for localized prostate cancer. N Engl J Med, 2011. 365(2): p. 107–18.

5. Spratt, D.E., et al., Androgen Receptor Upregulation Mediates Radioresistance after Ionizing Radiation. Cancer Res, 2015. 75(22): p. 4688–96.

6. Devos, G., et al., Neoadjuvant hormonal therapy before radical prostatectomy in high-risk prostate cancer. Nat Rev Urol, 2021. 18(12): p. 739–762.

7. Krafft, U., et al., A New Chapter in Neoadjuvant Therapy for High-risk Prostate Cancer? Eur Urol, 2023.

8. Karzai, F., et al., Sequential Prostate Magnetic Resonance Imaging in Newly Diagnosed High-risk Prostate Cancer Treated with Neoadjuvant Enzalutamide is Predictive of Therapeutic Response. Clin Cancer Res, 2021. 27(2): p. 429–437.

9. McKay, R.R., et al., Results of a Randomized Phase II Trial of Intense Androgen Deprivation Therapy prior to Radical Prostatectomy in Men with High-Risk Localized Prostate Cancer. J Urol, 2021: p. 101097JU0000000000001702.

10. McKay, R.R., et al., Evaluation of Intense Androgen Deprivation Before Prostatectomy: A Randomized Phase II Trial of Enzalutamide and Leuprolide With or Without Abiraterone. J Clin Oncol, 2019. 37(11): p. 923–931.

11. Montgomery, B., et al., Neoadjuvant Enzalutamide Prior to Prostatectomy. Clin Cancer Res, 2017. 23(9): p. 2169–2176.

12. Taplin, M.E., et al., Intense androgen-deprivation therapy with abiraterone acetate plus leuprolide acetate in patients with localized high-risk prostate cancer: results of a randomized phase II neoadjuvant study. J Clin Oncol, 2014. 32(33): p. 3705–15.

13. Devos, G., et al., ARNEO: A Randomized Phase II Trial of Neoadjuvant Degarelix with or Without Apalutamide Prior to Radical Prostatectomy for High-risk Prostate Cancer. Eur Urol, 2023. 83(6): p. 508–518.

14. Fizazi, K., et al., Androgen deprivation therapy plus docetaxel and estramustine versus androgen deprivation therapy alone for high-risk localised prostate cancer (GETUG 12): a phase 3 randomised controlled trial. Lancet Oncol, 2015. 16(7): p. 787–94.

15. Eastham, J.A., et al., Cancer and Leukemia Group B 90203 (Alliance): Radical Prostatectomy With or Without Neoadjuvant Chemohormonal Therapy in Localized, High-Risk Prostate Cancer. J Clin Oncol, 2020. 38(26): p. 3042–3050.

16. Febbo, P.G., et al., Neoadjuvant docetaxel before radical prostatectomy in patients with high-risk localized prostate cancer. Clin Cancer Res, 2005. 11(14): p. 5233–40.

17. Shenderov, E., et al., Neoadjuvant enoblituzumab in localized prostate cancer: a single-arm, phase 2 trial. Nat Med, 2023. 29(4): p. 888–897.

18. Abdul Sater, H., et al., Neoadjuvant PROSTVAC prior to radical prostatectomy enhances T-cell infiltration into the tumor immune microenvironment in men with prostate cancer. J Immunother Cancer, 2020. 8(1).

19. Vuky, J., et al., Phase II trial of neoadjuvant docetaxel and CG1940/CG8711 followed by radical prostatectomy in patients with high-risk clinically localized prostate cancer. Oncologist, 2013. 18(6): p. 687–8.

20. McKay, R.R., et al., Outcomes Post Neoadjuvant Intense Hormone Therapy and Surgery for Patients with High-Risk Localized Prostate Cancer: Results of a Pooled Analysis of Contemporary Clinical Trials. J Urol, 2021: p. 101097JU0000000000001632.

21. McKay, R.R., et al., Post prostatectomy outcomes of patients with high-risk prostate cancer treated with neoadjuvant androgen blockade. Prostate Cancer Prostatic Dis, 2018. 21(3): p. 364–372.

22. Wilkinson, S., et al., Nascent Prostate Cancer Heterogeneity Drives Evolution and Resistance to Intense Hormonal Therapy. Eur Urol, 2021.

23. Wilkinson, S., et al., A case report of multiple primary prostate tumors with differential drug sensitivity. Nat Commun, 2020. 11(1): p. 837.

24. Tewari, A.K., et al., Molecular features of exceptional response to neoadjuvant anti-androgen therapy in high-risk localized prostate cancer. Cell Rep, 2021. 36(10): p. 109665.

25. Beltran, H., et al., Impact of Therapy on Genomics and Transcriptomics in High-Risk Prostate Cancer Treated with Neoadjuvant Docetaxel and Androgen Deprivation Therapy. Clin Cancer Res, 2017. 23(22): p. 6802–6811.

26. Ramos, P. and M. Bentires-Alj, Mechanism-based cancer therapy: resistance to therapy, therapy for resistance. Oncogene, 2015. 34(28): p. 3617–26.

27. Robinson, D., et al., Integrative Clinical Genomics of Advanced Prostate Cancer. Cell, 2015. 161(5): p. 1215–1228.

28. Yu, Z., et al., Rapid Induction of Androgen Receptor Splice Variants by Androgen Deprivation in Prostate Cancer. Clinical Cancer Research, 2014. 20(6): p. 1590–1600.

29. Abida, W., et al., Genomic correlates of clinical outcome in advanced prostate cancer. Proc Natl Acad Sci U S A, 2019. 116(23): p. 11428–11436.

30. Alumkal, J.J., et al., Transcriptional profiling identifies an androgen receptor activity-low, stemness program associated with enzalutamide resistance. Proc Natl Acad Sci U S A, 2020. 117(22): p. 12315–12323.

31. Spratt, D.E., et al., Transcriptomic Heterogeneity of Androgen Receptor Activity Defines a de novo low AR-Active Subclass in Treatment Naive Primary Prostate Cancer. Clin Cancer Res, 2019. 25(22): p. 6721–6730.

32. Zhang, Z., et al., Tumor Microenvironment-Derived NRG1 Promotes Antiandrogen Resistance in Prostate Cancer. Cancer Cell, 2020. 38(2): p. 279–296 e9.

33. Sowalsky, A.G., et al., Neoadjuvant-Intensive Androgen Deprivation Therapy Selects for Prostate Tumor Foci with Diverse Subclonal Oncogenic Alterations. Cancer Res, 2018. 78(16): p. 4716–4730.

34. Cai, C., et al., Androgen receptor expression in prostate cancer cells is suppressed by activation of epidermal growth factor receptor and ErbB2. Cancer Res, 2009. 69(12): p. 5202–9.

35. Gao, S., et al., ErbB2 Signaling Increases Androgen Receptor Expression in Abiraterone-Resistant Prostate Cancer. Clin Cancer Res, 2016. 22(14): p. 3672–82.

36. Mellinghoff, I.K., et al., HER2/neu kinase-dependent modulation of androgen receptor function through effects on DNA binding and stability. Cancer Cell, 2004. 6(5): p. 517–27.

37. Signoretti, S., et al., Her-2-neu expression and progression toward androgen independence in human prostate cancer. J Natl Cancer Inst, 2000. 92(23): p. 1918–25.

38. Lundberg, A., et al., The Genomic and Epigenomic Landscape of Double-Negative Metastatic Prostate Cancer. Cancer Res, 2023. 83(16): p. 2763–2774.

39. Penning, T.M., AKR1C3 (type 5 17beta-hydroxysteroid dehydrogenase/prostaglandin F synthase): Roles in malignancy and endocrine disorders. Mol Cell Endocrinol, 2019. 489: p. 82–91.

40. Ku, A.T., S. Wilkinson, and A.G. Sowalsky, Comparison of approaches to transcriptomic analysis in multi-sampled tumors. Brief Bioinform, 2021. 22(6).

41. Craft, N., et al., A mechanism for hormone-independent prostate cancer through modulation of androgen receptor signaling by the HER-2/neu tyrosine kinase. Nat Med, 1999. 5(3): p. 280–5.

42. Cancer Genome Atlas Research Network, The Molecular Taxonomy of Primary Prostate Cancer. Cell, 2015. 163(4): p. 1011–25.

43. Taavitsainen, S., et al., Single-cell ATAC and RNA sequencing reveal pre-existing and persistent cells associated with prostate cancer relapse. Nat Commun, 2021. 12(1): p. 5307.

44. Morris, M.J., et al., HER-2 profiling and targeting in prostate carcinoma. Cancer, 2002. 94(4): p. 980–6.

45. Osman, I., et al., HER-2/neu (p185neu) protein expression in the natural or treated history of prostate cancer. Clin Cancer Res, 2001. 7(9): p. 2643–7.

46. Takeda, D.Y., et al., A Somatically Acquired Enhancer of the Androgen Receptor Is a Noncoding Driver in Advanced Prostate Cancer. Cell, 2018. 174(2): p. 422–432 e13.

47. Quigley, D.A., et al., Genomic Hallmarks and Structural Variation in Metastatic Prostate Cancer. Cell, 2018. 174(3): p. 758–769 e9.

48. Ziada, A., et al., The use of trastuzumab in the treatment of hormone refractory prostate cancer; phase II trial. Prostate, 2004. 60(4): p. 332–7.

49. Estephan, F., et al., The prevalence and clinical significance of HER2 expression in prostate adenocarcinoma. Annals of Diagnostic Pathology, 2023. 67: p. 152219.

50. Minner, S., et al., Low level HER2 overexpression is associated with rapid tumor cell proliferation and poor prognosis in prostate cancer. Clin Cancer Res, 2010. 16(5): p. 1553–60.

51. Ahmad, I., et al., HER2 overcomes PTEN (loss)-induced senescence to cause aggressive prostate cancer. Proc Natl Acad Sci U S A, 2011. 108(39): p. 16392–7.

52. Smith, M.R., et al., Darolutamide and Survival in Metastatic, Hormone-Sensitive Prostate Cancer. N Engl J Med, 2022. 386(12): p. 1132–1142.

53. Davis, I.D., et al., Enzalutamide with Standard First-Line Therapy in Metastatic Prostate Cancer. N Engl J Med, 2019. 381(2): p. 121–131.

54. Dalin, M.G., et al., Comprehensive Molecular Characterization of Salivary Duct Carcinoma Reveals Actionable Targets and Similarity to Apocrine Breast Cancer. Clin Cancer Res, 2016. 22(18): p. 4623–33.

55. McAfee, J.L., et al., ERBB2 Amplification and HER2 Expression in Salivary Duct Carcinoma: Evaluation of Scoring Guidelines and Potential for Expanded Anti-HER2 Therapy. Mod Pathol, 2023. 36(10): p. 100273.

56. Sowalsky, A.G., et al., Assessment of Androgen Receptor Splice Variant-7 as a Biomarker of Clinical Response in Castration-Sensitive Prostate Cancer. Clin Cancer Res, 2022. 28(16): p. 3509–3525.

